# Latent Blowout of COVID-19 Globally: An Effort to Healthcare Alertness via Medical GIS Approach

**DOI:** 10.1101/2020.04.27.20082503

**Authors:** Laxmi Kant Sharma, Rajani Kant Verma

**Affiliations:** Department of Environmental Science, School of Earth Sciences, Central University of Rajasthan, Bandarsindri-305817, Ajmer (Rajasthan), India

## Abstract

Since January 2020, the COVID-19 pandemic has been escalating from North America to Asia. Various studies projected the spread of pandemic globally, using air passenger data from an infected area. But there could be various parameters that can be the basis for the forecasting of the pandemic. Current research adopts the Medical GIS approach and incorporates critical parameters from various domains to create a global alertness scale to combat the pandemic. The finding of the study ranks the countries on a 1 to 9 scale based on the spatial alertness In this context, the study focuses on the role of GIS techniques as an enabler to fight against the global pandemic and could be beneficial for the authorities to adopt timely preventive actions.

**One Sentence Summary:** Global Alertness Ranking to combat COVID-19

---

The worldwide spread of epidemics is a complex and network-driven phenomenon. The multiscale epidemic environment makes it challenging to develop systematic pathways to combat them. It also makes it difficult to identify/ predict the external factors responsible for the rapid outbreak of epidemics in due course of time and locate its sources. Wuhan (China) had reported an outburst of uncommon pneumonia since December 31, 2019, which was instigated by COVID-19, a novel coronavirus as defined by WHO. The city converted into an epicenter of an infectious disease with unknown origin and rapid, life-threatening symptoms by the last week of January 2020. This sudden outbreak raised deep concern amongst the health authorities in China. However, the delayed responses lead to worsening the situation and increased the number of infected cases and heighten the death toll in China. It was due to the delayed preliminary screening of contacts, epidemiological data collection, and the development of diagnostic kits by the authorities. Further, a global outburst of this threatening disease occurred through the dissemination of infected cases from China to different parts of the world. In the first week of February 2020, WHO declared COVID-19 as a pandemic. The cases have been intensifying in North America, the Middle East, Europe, and Asia. The number of cases of pandemic outside mainland China had increased drastically by the first week of March 2020 especially in, Italy, Spain, and France, and now cases of COVID-19 are no longer limited to one country. Thousands of instances of disseminated contagion as well as deaths have reported in the whole world till date, and further disseminating through person to person contact. Chan et al. *(8)*, in The Lancet recently said a rapid increase of COVID 19 cases in the healthcare staff, indicating the severeness of human to human transmission of this disease. Huang et al. *(18)* reported clinical symptoms of the newly admitted patients from the selected hospitals in china, and the outcomes provide data regarding the severity of the COVID-19 epidemic infections. Results emphasized the following preliminary symptoms viz. dry cough, fever, and difficulty in breathing. Chan et al. *(8)* described that this novel zoonotic viral pathogen adapts more quickly when transmitted from human to human, thus intensifying the spread of this disease. Figure 1 reports the severity of the pandemic based on the data collected from the dashboard of the Centre for Systems Science and Engineering (CSSE) at Johns Hopkins University of Medicine *(12)*. The total confirmed global cases of COVID-19 as on March 30, 2020, were 723,700, whereas the total deaths were 34,005. However, 152,032 cases reported to recovered consequently *(12,33,34)*.

**Fig. 1.**
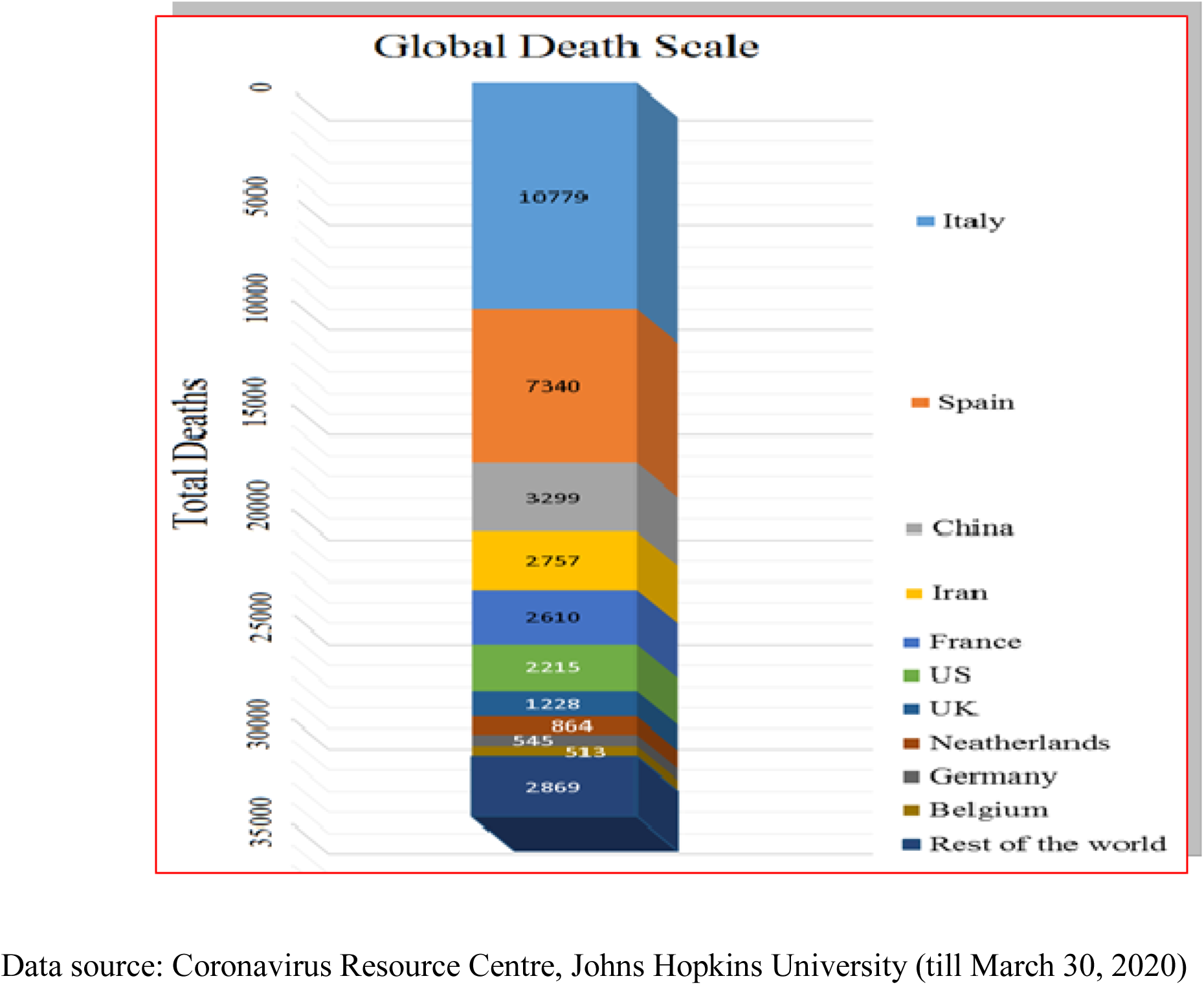
Country-wise death reported on a global scale to COVID-19.

Epidemiology is a stream of medical science dealing with the spreading, prevention, control, and suppression of infectious diseases in the human community. Human actions act as triggers for the spread of epidemics and contribute to foremost factors like climate change, environmental hazards, political, socio-economic, disease patterns, and stimulus emergence. Over the past two decades, there has been a sharp increase in the number of epidemiological studies engaging GIS, particularly in the domain of health inconsistency, resource availability, and health-related performance *(11)*. Further there has been a persistent use of GIS in introductory arenas of malignancy and environmental epidemiology *(14,29)*. GIS has also been used broadly in epidemiology for disease surveillance and observing intermediation *(12)*. In addition to traditional epidemiological approach, medical geographers can measure the spatial distribution and accessibility of health facilities using GIS *(10,15,16)*. Since the 1990s Medical Geographic Information Systems (Medical GIS) has become extremely valuable and is being applied as public health reformist system. It is used for identifying and investigating medical and epidemiological indicators for diseases ranging from cholera to cancer *(5,14,15,28)*. The most extensively used tools for supporting this system are dynamic continuous area space-time analysis, spatial statistics, agent-based modeling, cellular automata, geographical analysis machine, self-organizing maps etc. *(14,15,21)*. Medical GIS technology offers exceptional means for visualizing epidemiological data, revealing disease trends and their inter-relationships *(21,35)*. It integrates the spatial and non-spatial data for analysis and decision making. It generates a widespread response, thereby reducing the health hazards in the population, during, before, and after the violent events, from local to the global level *(14)*. Medical GIS serves for multi-disease surveillance activities using different operations viz. buffer analysis; network analysis; statistical analysis; time series analysis and spatio-temporal analysis to identify the health centers, movement of carriers, the intensity of spread etc. *(4,22)*. Operative response provided by these analyses requires a commitment to risk reduction, prevention, control, and timely financial support. It is proved that Medical GIS approach is playing an important role for advance planning in health education and emergency health care provisions leading to infrastructure protection and reduction in financial burden. The current study explores the Medical GIS approach in identifying the prone area of the disease using GIS overlay analysis for vulnerable groups on a global scale. The task of expanding worldwide health care disease mapping using GIS has tremendous positive effects. This automated system for managing spatial data permits an informed user to choose between options when geographic dissemination is part of the problem *(1,2)*. Recently Joseph et al. *(20)* studied the worldwide spread of COVID-19, from Wuhan and adjacent towns. They forecasted the national and global spread of 2019-nCoV using a single parameter approach. They used periodic flight reservations over the period December 31, 2019, to January 28, 2020, and used the Markov Chain Monte Carlo method to forecast the corona spread. Their findings suggest that independent, self-sustaining human-to-human spread has already started, and this has resulted in a global outburst of the disease as major Chinese cities are global transport hubs. The current study estimates five parameters for analysis of the global latent spread of COVID-19 from different available sources as an extension of the previous study *(20)*. Data were divided into five categories viz. the total number of infected cases country-wise; country-wise air passenger's pressure from top ten busy airports of china including Wuhan; world population density, world meat consumption; world health ranking (country-wise). Entire data parameters converted into spatial domains and assigned weightage based on GIS fuzzy overlay function. Results represent highest to lowest health care alertness globally, on a scale of 1 to 9. As the study based on global scale parameters, hence the results may vary from local to a global level. Countries across the world are taking necessary actions to prevent COVID-19 pandemic using alternate measures and techniques. In this context, the current study focuses on the role of Medical GIS techniques as an enabler to fight against global pandemic diseases.

## Materials and Methods

### Data description

Our study collects data on five parameters for analysis of the global latent spread of COVID-19 from different sources. Data divided into two categories first is core data, and the second is subsidiary data. Core data compiled with two parameters in which one is the total number of coronavirus infected cases according to country wise and second is country-wise air passenger's pressure coming from the top ten busy airports of china including Wuhan. Subsidiary data comprises three parameters. The first parameter is the country-wise population density. Population density is an essential parameter for epidemiology. High population density will show a high risk of transmission of disease. According to Merler and Ajelli *(27)*, influenza activity is affected by the spatial heterogeneity of population density-epidemic reaches much earlier than rural areas. Countries that have large household groups and large fractions of students in the population would face more severe outbreaks *(9,27)*. So, we take population density as a first subsidiary parameter. The second parameter consists of country-wise meat consumption. Researchers confirm that coronavirus is a zoonotic virus, so according to Duizer and Koopmans13 when the emergence of CoV is reported or noticed, foodborne transmission should be considered to determine the development and spread of the virus and putting countermeasures into place *(13)*. The contaminations by live animals or rodents are a possibility because sold animals usually come from various sources in different vendors *(13,19)*. The cause may be apparent in the future, but now we take total meat consumption per person as a second subsidiary parameter. The third parameter is the country-wise health care ranking. Country planning, preparedness, and health facilities may play a vital role in preventing an epidemic. It believed that the government of any country would be prepared to act rapidly and effectively to combat an outbreak according to their health facilities. So, we take the health care rank of countries across the globe as a third subsidiary parameter.

All data parameters converted into the spatial domain. Subsequently, the weighted overlay GIS function applied to the fuzzy overlay technique used with a sum operator for data normalization of the subsidiary data layer. Further, the output reclassified into nine classes subsequently, weights assigned for core data based on the passenger pressure and the number of infected cases. Hence three layers were prepared, namely, air passenger, the number of infected cases, and reclassified segment of normalized secondary data. After the preparation of these layers, the fuzzy overlay function applied to these layers with the sum operator as denoted in Figure S1.

## Weighted Overlay

A weighted overlay is a technique in which we applied scale values for different and disparate inputs to produce an integrated investigation. Based on application and suitability in a single raster, the prerequisite has to state the weightage to the particular classes. The weighted overlay matrix provides a 1 to 10 scale to assign the weightage to an individual input raster. The more commendatory class for each input standard will reclassify on the higher values as per Baidya et al. *(6)*.

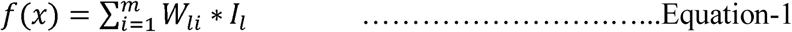

Source: Baidya et al. *(6)*

Here,

*m* is the number of input raster,
*W_li_* is the weightage value of an *i*th pixel of lth layer,
*I_l_* is the percentage influence of the lth layer.

In this equation, layers multiplied as per suitable weightage. For each cell, the corresponding values added together. Weighted overlay estimates that highly suitable factors consequence for the higher costs in the output raster.

### Fuzzy overlay analysis

Fuzzy overlay approach is a mathematical function based on set theory representing multivalued logic that approaches with uncertainty, where a set correlates to a class as per Adnam et al. *(1)*, Aleksa et al. *(3)*, Cakıt and Karwowski *(7,23)*. It convert the data values to a standard scale, but the changed values signify the probability to belonging in a specified class. Fuzzy overlay approach follows the principle of fuzzy logic to explain traditional overlay analysis in GIS-based problems. It includes several operators such as fuzzy SUM, fuzzy OR, fuzzy AND, fuzzy GAMMA, and fuzzy product that provides as a different characteristic of each class to the multiple input measures by Kirschbaum et al. *(23)*. The current study uses a fuzzy SUM operator.

**Fig. S1.**
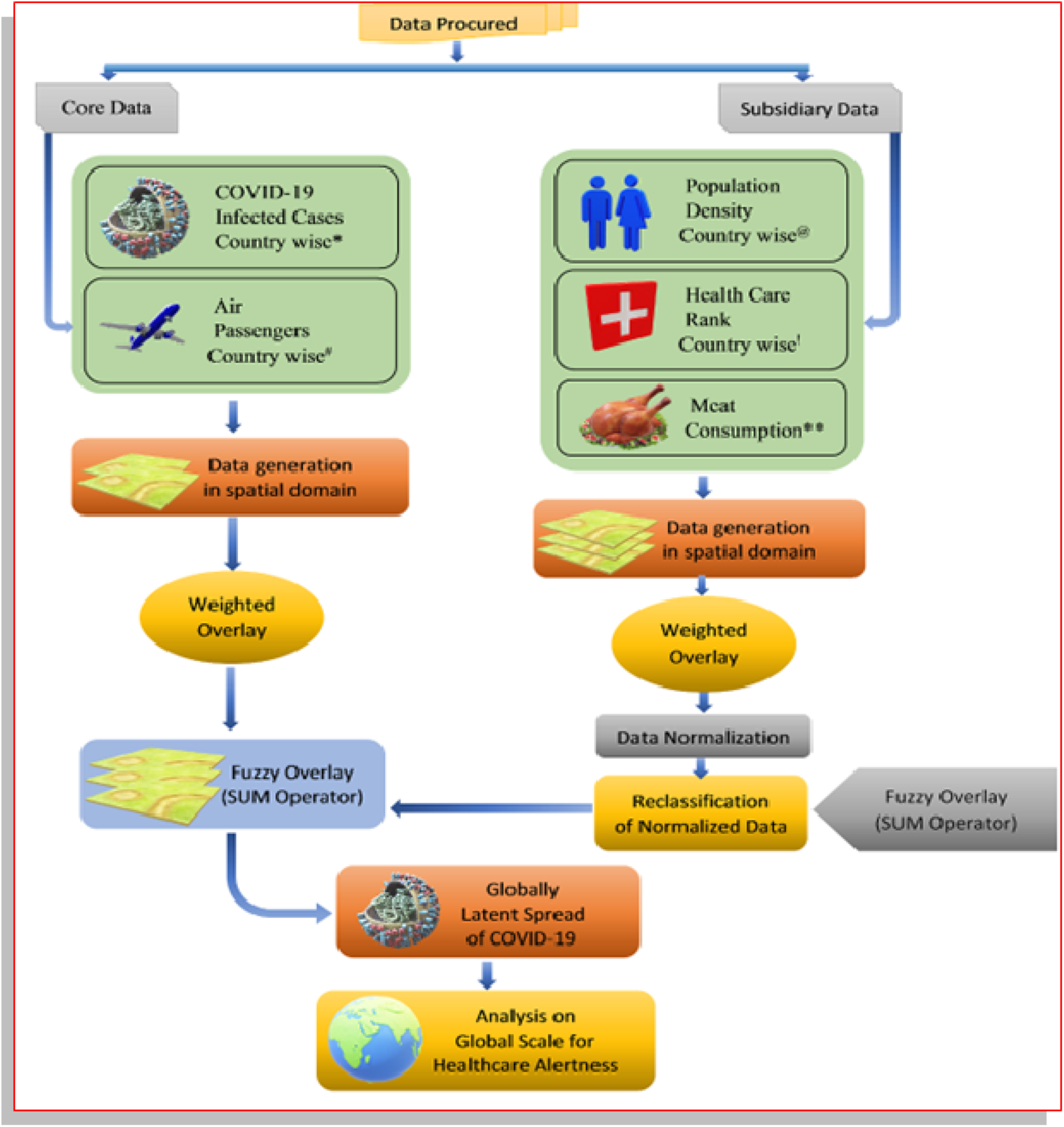
Overall, an applied methodology for the latent spread of COVID-19 globally **Data Sources** **^*^Coronavirus Resource Center** https://coronavirus.jhu.edu/map.html *(12)* (Reported by March 4, 2020) **^#^Air Passengers Data source:** Mao et al. *(25)* **^@^Population Density Data Source:** http://worldpopulationreview.com/ *(30)* **^!^Health Care Data Source:** http://worldpopulationreview.com/countries/best-healthcare-in-the-world/ *(17)* **^**^Meat Consumption Data Source**: https://www.scribd.com/doc/91840616/Meat-Consumption-Per-Person *(26)*

**Fig. S2.**
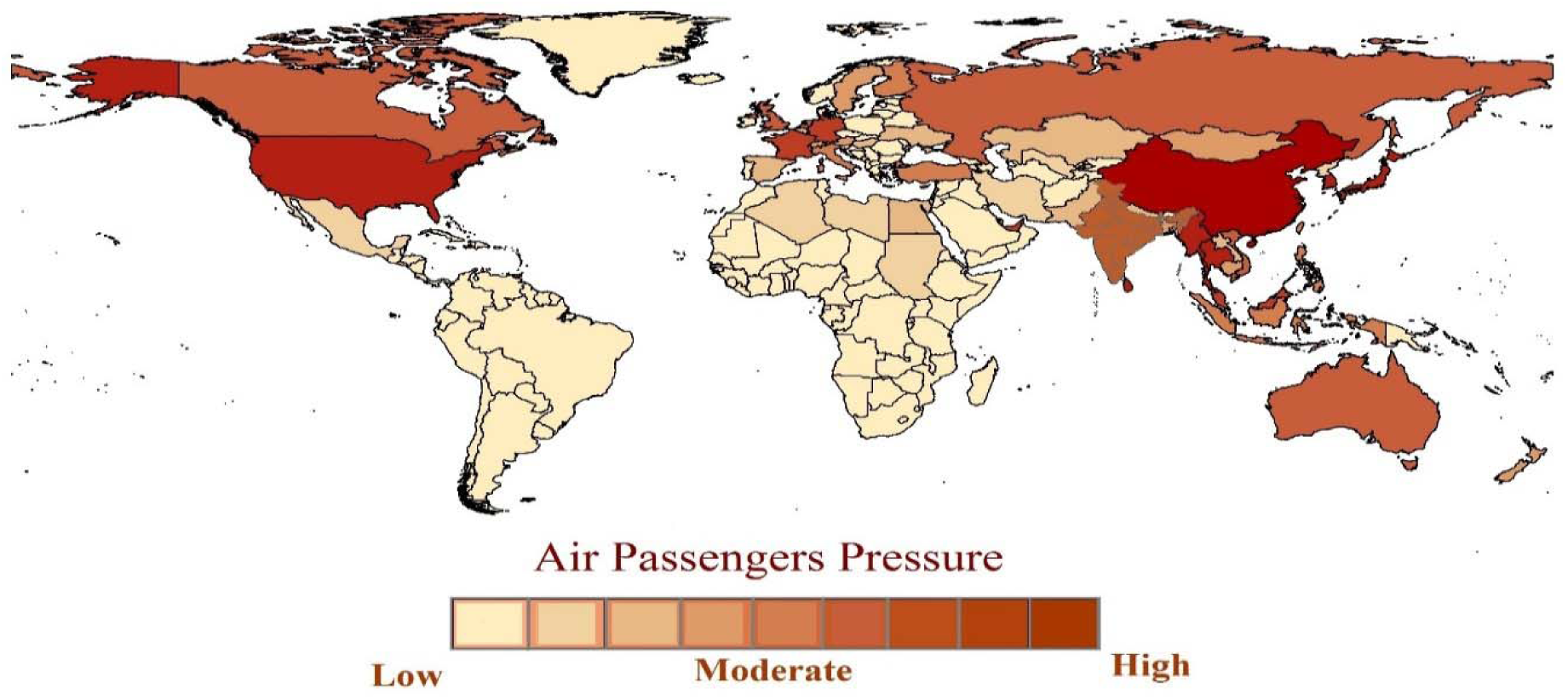
Air passengers pressure data based on top ten air connectivity routes from china to rest of the world

**Fig. S3.**
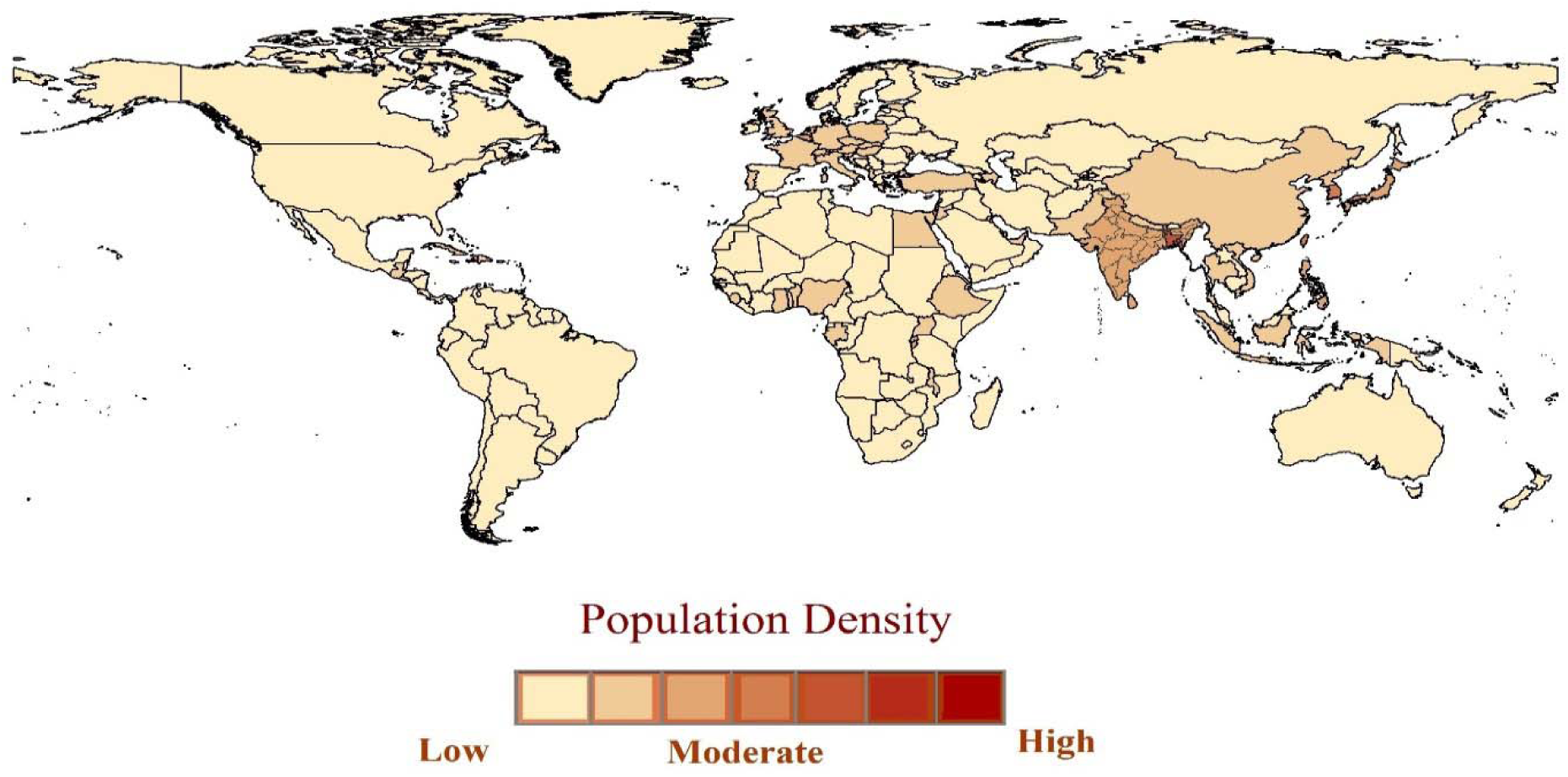
Country wise spatial distribution of population density

**Fig. S4.**
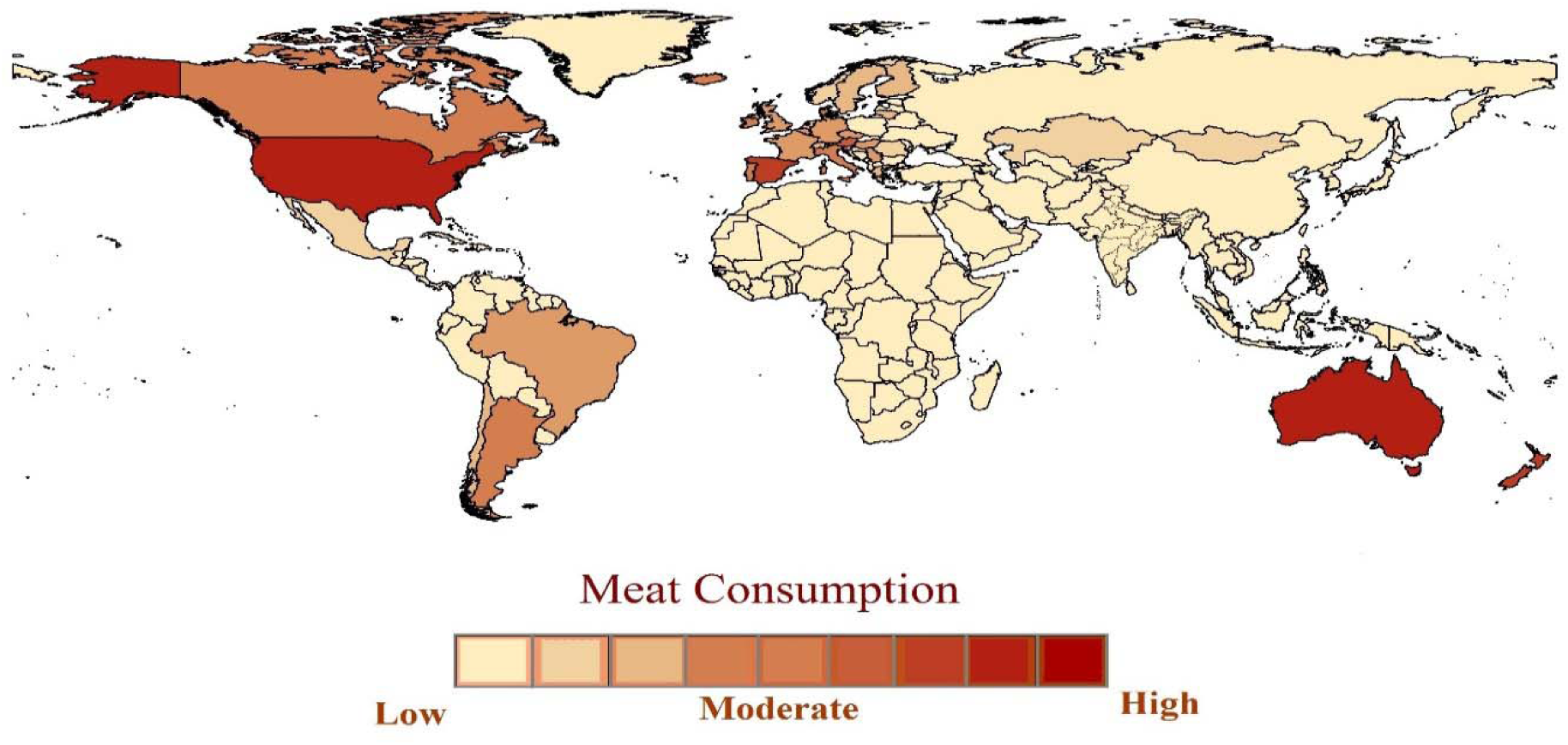
Country wise spatial distribution of meat consumption

**Fig. S5.**
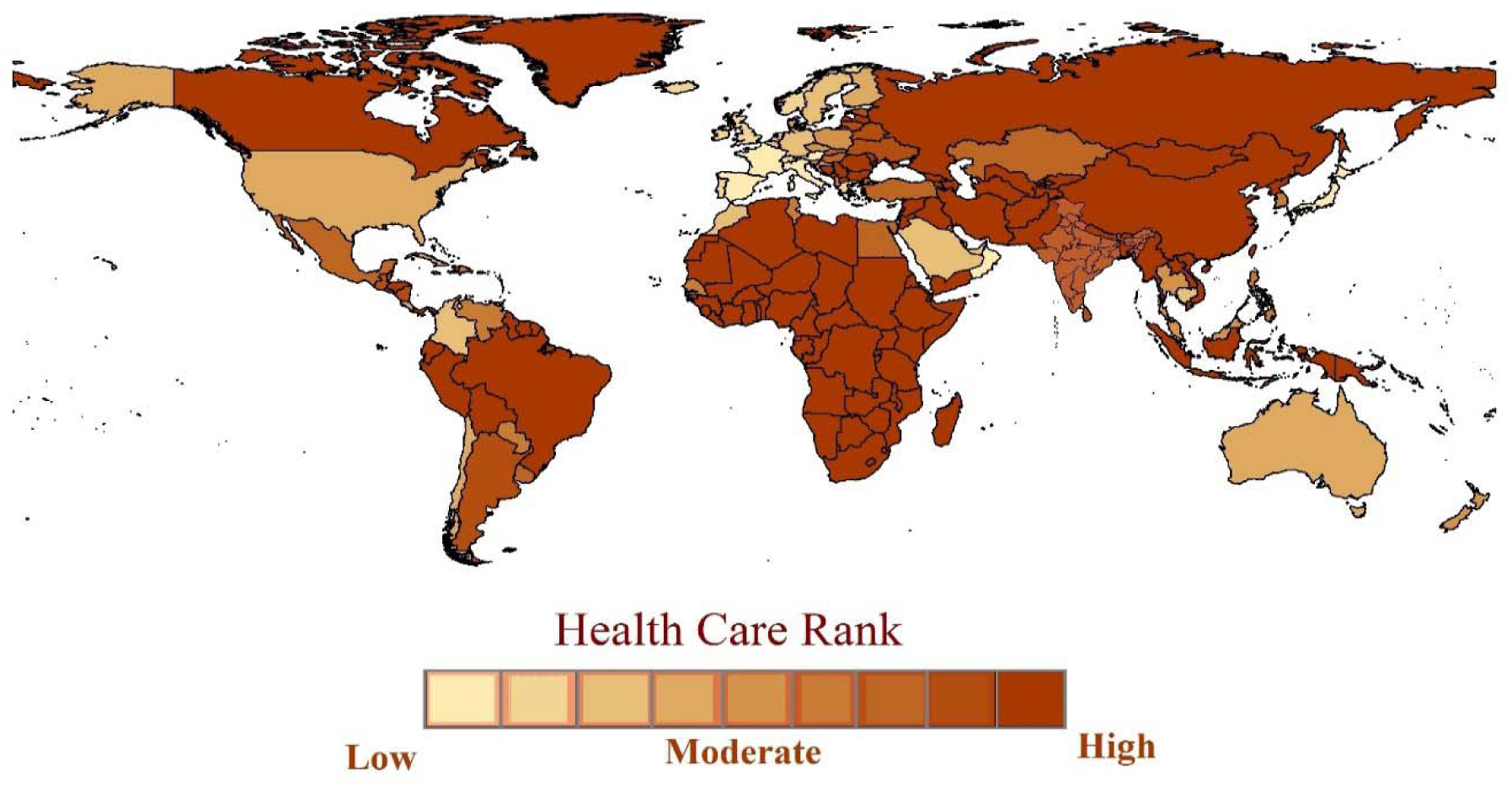
Country wise spatial distribution of health care

## Results

The finding of the study ranks the countries based on the spatial alertness regarding the spread of COVID 19 on a scale of 1 to 9. Where 1 indicates the highest alertness, and 9 shows the lowest alertness. The study uses a GIS overlay model and incorporates both weighted overlay and fuzzy overlay functions. Table 1 indicates the worldwide Ranking of alertness for COVID-19 for 102 countries based on five parameters. In rank 1, criterion countries are at high risk and prone to significant outbursts of the pandemic. This criterion has 28 countries, including China, Japan, South Korea, Iran, Greece, Italy, Germany, Netherlands, France, Spain, the UK, Canada, US, and Australia. Recent statistics reporting the deaths by pandemic (figure 1) supports the results of our study, confirming the outbreak in Italy, Spain, France, Germany, Iran, the UK, and the USA.

**Table 1.**
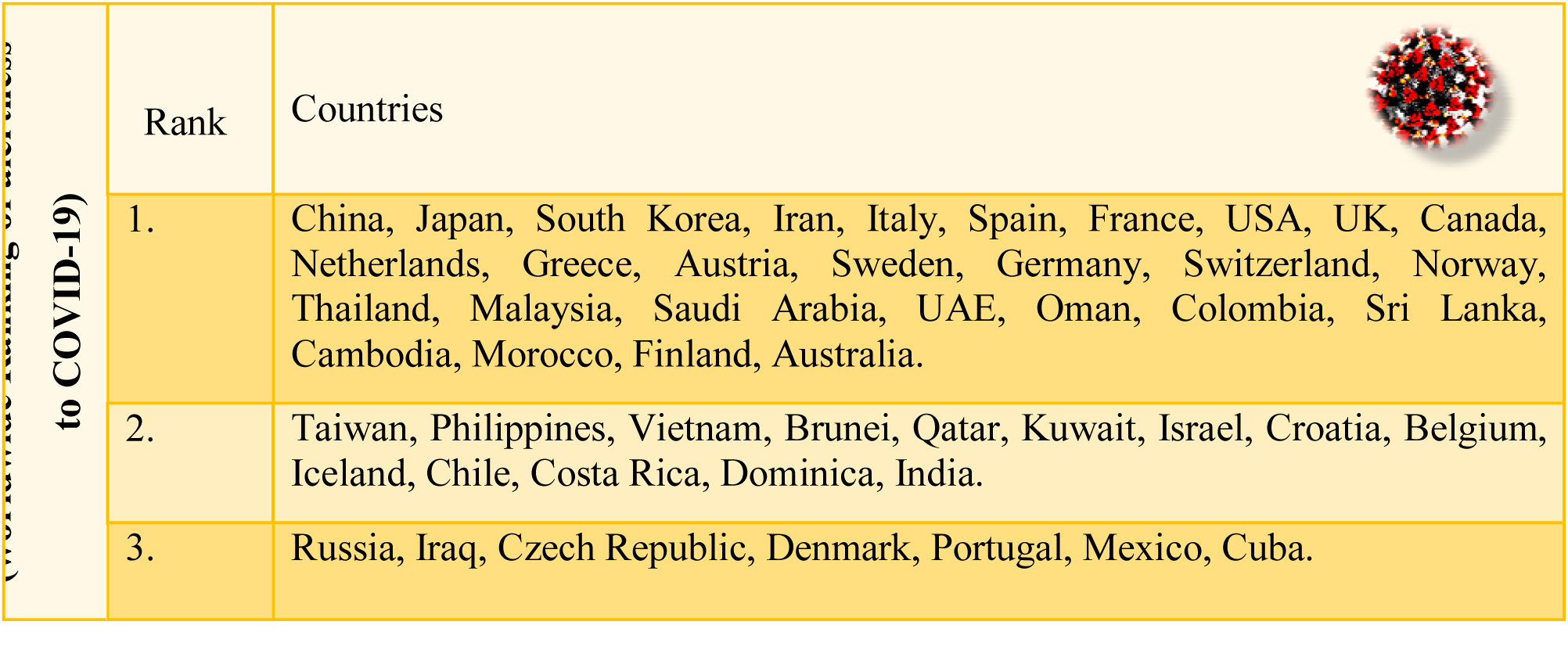
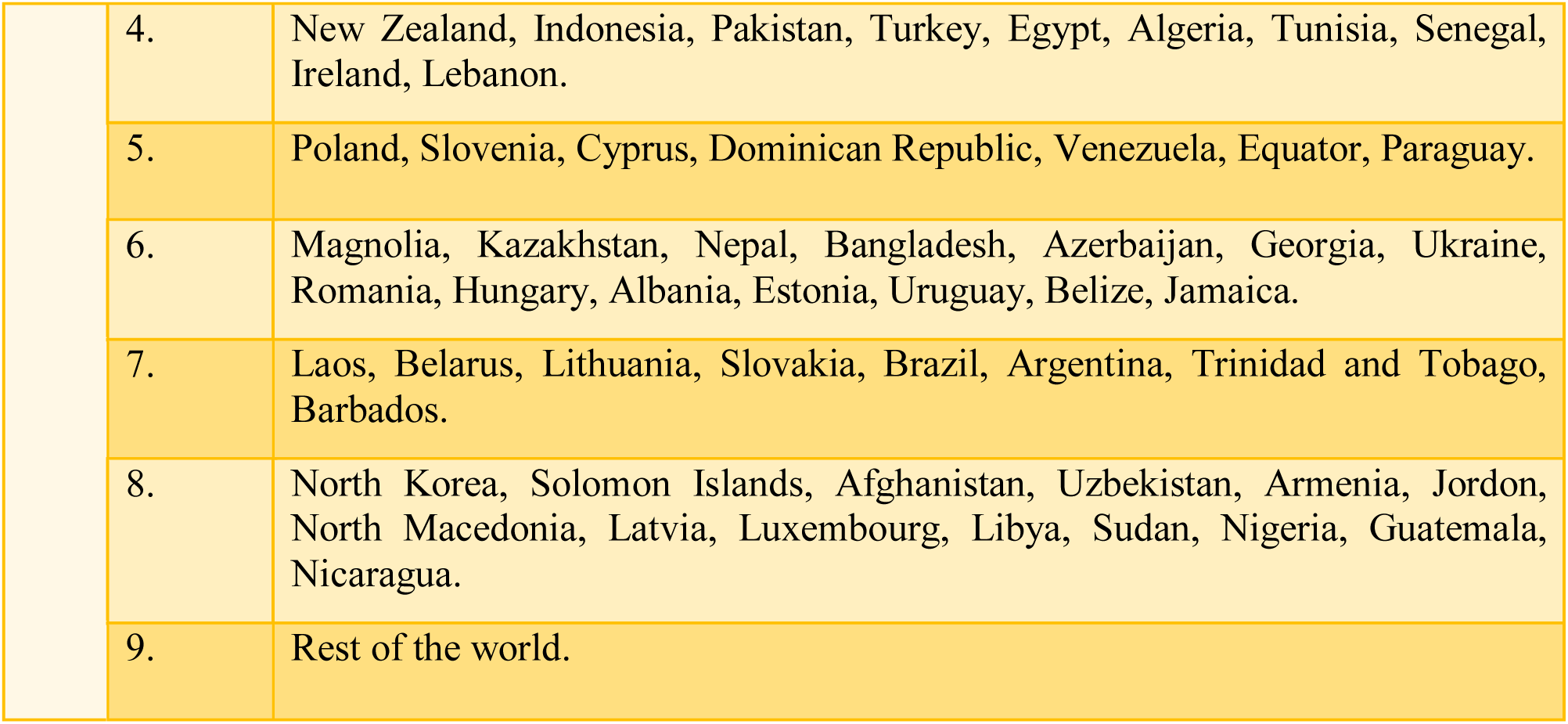
List of countries according to alertness based on five parameters.

| to COVID-19) | Rank | Countries |
| --- | --- | --- |
|  | 1. | China, Japan, South Korea, Iran, Italy, Spain, France, USA, UK, Canada, Netherlands, Greece, Austria, Sweden, Germany, Switzerland, Norway, Thailand, Malaysia, Saudi Arabia, UAE, Oman, Colombia, Sri Lanka, Cambodia, Morocco, Finland, Australia. |
|  | 2. | Taiwan, Philippines, Vietnam, Brunei, Qatar, Kuwait, Israel, Croatia, Belgium, Iceland, Chile, Costa Rica, Dominica, India. |
|  | 3. | Russia, Iraq, Czech Republic, Denmark, Portugal, Mexico, Cuba. |

|  |  |
| --- | --- |
| 4. | New Zealand, Indonesia, Pakistan, Turkey, Egypt, Algeria, Tunisia, Senegal, Ireland, Lebanon. |
| 5. | Poland, Slovenia, Cyprus, Dominican Republic, Venezuela, Equator, Paraguay. |
| 6. | Magnolia, Kazakhstan, Nepal, Bangladesh, Azerbaijan, Georgia, Ukraine, Romania, Hungary, Albania, Estonia, Uruguay, Belize, Jamaica. |
| 7. | Laos, Belarus, Lithuania, Slovakia, Brazil, Argentina, Trinidad and Tobago, Barbados. |
| 8. | North Korea, Solomon Islands, Afghanistan, Uzbekistan, Armenia, Jordon, North Macedonia, Latvia, Luxembourg, Libya, Sudan, Nigeria, Guatemala, Nicaragua. |
| 9. | Rest of the world. |

14 countries are falling in the rank 2 criteria majorly constituting Asian and Middle East countries. As these countries are on the verge of the outburst of the epidemic, our study could be beneficial for the authorities to adopt timely preventive actions like social distancing, limited international and domestic travels, self-isolation, public awareness drives, increasing the healthcare capacities, face masks and PPE kits for the medical practitioners and staff. The countries falling in the range from 3 to 9 should also start preparing a systematic control mechanism and learn from the mistakes of the countries where the situation has worsened. As WHO suggests that to date, no vaccine is available to cure this epidemic, so protection is the only cure to combat COVID 19. In such a scenario, our study could be a guiding force for practitioners, authorities, and countries which are fighting hard to stop this epidemic. Figure 2 represents the worldwide spatial distribution of alertness to COVID-19 on the world map based on the medical GIS technique.

**Fig. 2.**
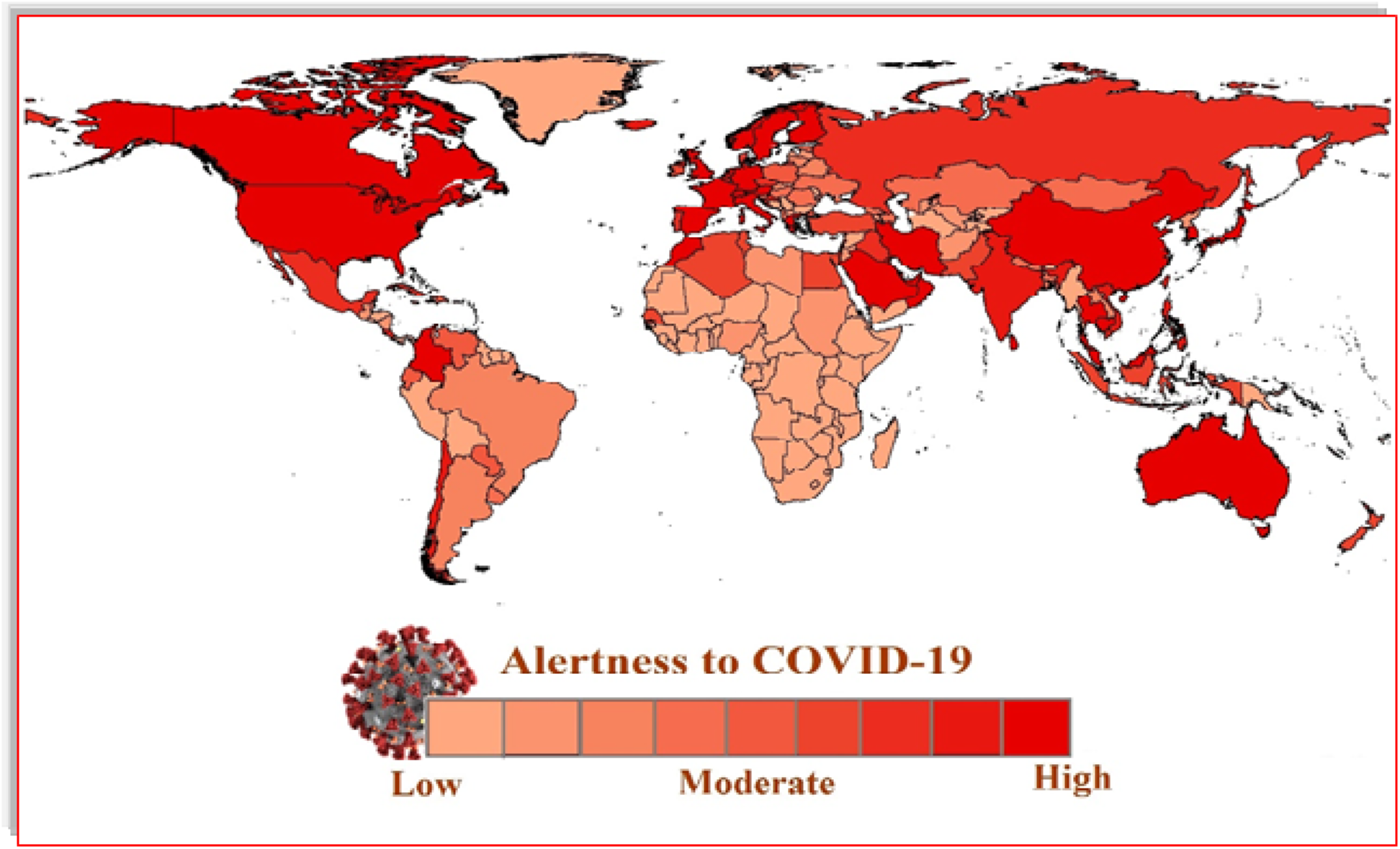
Worldwide spatial distribution of alertness to COVID-19.

As a robustness check, we estimated the relationship between the results of alertness and the number of reported deaths till March 30, 2020. Figure 3 graphically indicates a linear relationship between the two variables and thus comprehends the effectiveness of results.

**Fig. 3.**
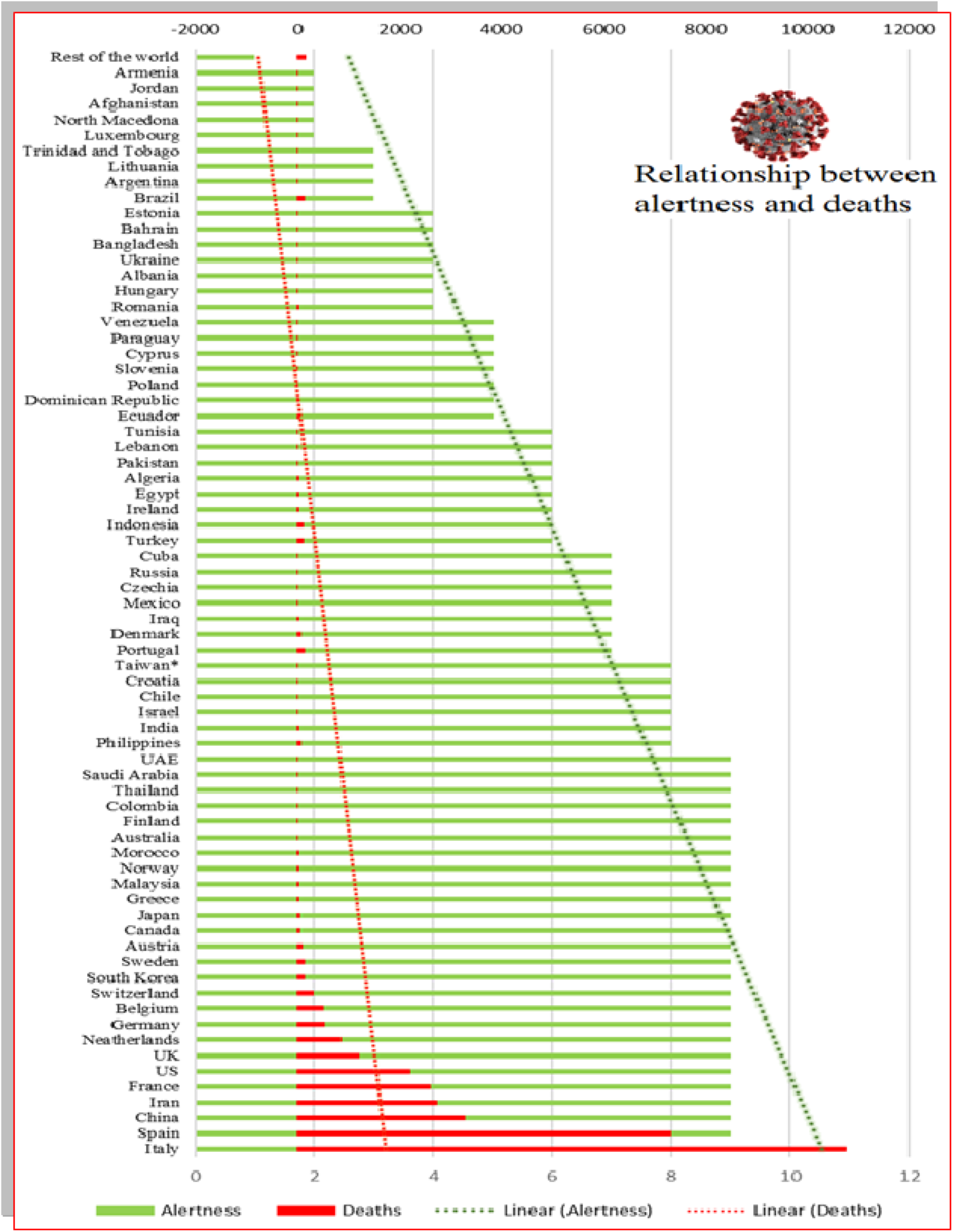
Relationship between alertness and death reported. (till March 30, 2020)

## Discussion

COVID-19 is an infectious disease spread through a new virus, characterized by cough, strong infectivity, fever, elongated incubation period, and difficulty of detection, which has led to the sudden outbreak and the rapid development of a pandemic. The present situation requires medical GIS data to generate quick responses and investigations for the timely prevention and control of the disease. This requires a rapid source of information about the study of epidemic dynamics. In this study, we analyzed the spatial Ranking of global alertness scale against the spread of COVID 19, based on various parameters related either directly or indirectly, which previous studies ignored. The study focused on five parameters from different dimensions. These parameters further characterized under two categories as core and subsidiary. Core data constituted a total number of coronavirus infected cases and country-wise air passenger's pressure. Subsidiary data formed country-wise population density, country-wise meat consumption, and country-wise health ranking. Based on them, a global scale geospatial index created, and 102 countries were ranked on a scale of 1 to 9 for alertness towards COVID 19 pandemic. Broader scale parameters used in the current study; hence, the results may fluctuate at the local to the global level. This study links GIS technology for the detection of disease in a pandemic situation. It establishes a strong bond between medical epidemiology and GIS systems to develop, identify, implement, and control mechanisms for efficient decision making for healthcare administrators and professionals. The ability of GIS to link disease information with spatial data makes it an asset in the progression of global healthcare systems *(31)*. The GIS overlay operations that we used in our research are almost parallel to those used by other scientists, who are working towards the same goal of characterizing the epidemic dynamics of COVID-19. The consensus on our approach provides some support for the validity of our predictions. An added strength of the current study is that the use of five crucial parameters that directly or indirectly influence the possible outbreak rating of pandemic COVID-19.

The current study provides broad directives on a global scale to combat the pandemic using an alertness scale; however, it has certain limitations. We used passenger travel data based on Mao Liang et al. *(25)* due to financial constraints. If real-time data would have used, the results could be more robust. We also assume that the pandemic spread of disease did not affect passenger's travel behavior. Further, our epidemic forecast based on one of the parameters, i.e., meat consumption data, which might not necessarily reflect the pandemic situation at a larger scale, especially in the occurrence of current public attentiveness and response concerning the health risk posed by of COVID-19. Further parameters related to seasonality, the origin of disease, accounting for airborne transmissibility, could be used as an extension of the study. Worldwide scientists and doctors are trying hard to develop a vaccine to cure the disease in the event of a second wave of infection. One of the current expansions in the fields of Medical GIS is coming from our growing ability to assemble mass amounts of information, known as 'Big Data' that can be helpful with determining urban mobility in case of an emergency evacuation *(24,32,36)*. During the period of a pandemic when the human-to-human transmission established and reported case numbers are rising exponentially, forecasting is of crucial importance for public health planning and control domestically and globally. Medical GIS could be used extensively with big data for strategically combating the pandemic by identifying those sites with the contiguous travel links with infected cities, quick deployment of preparedness plans, secured pharmaceutical supply chain, hospital supplies, and the necessary human resources to deal with the consequences of a global outbreak of this magnitude.

- One of the most influential aspects of this kind of study is its ability to recognize where epidemic diseases are most probably to spread next on a global scale by using the Medical GIS domain.
- By the spatial representation of infectious diseases in the GIS domain, we can analyze risk vulnerability for epidemics. We can make faster, better and necessary decisions in the field of public health and decision making before the outbreak.
- In epidemiology, it is necessary to understand an epidemic, its transmission factors, and its indicators on the GIS domain so that we will be able to understand how an epidemic spread through human to human and can analyze its spread rate and the correlation between its factors.
- The study may help in the monitoring and management of epidemics and health care alertness to save human lives.

## Data Availability

N/A

## Acknowledgments

Authors have acknowledged the uses of the DST-FIST supported lab for using GIS software’s.

## Funding

No funding source for this research.

## Author contributions

LKS: Designed the concept, analyzed data, interpreted the results, and wrote the manuscript.

RKV: Collected data and GIS work

## Competing interests

No competing interest for this research.

## Data and materials availability

We collated health data from publicly available data sources (news articles, press releases, and published reports from public health agencies, openly assessed web sources, etc.). All the information that we used documented in the article with proper citations.

## Notes

### Competing Interest Statement

The authors have declared no competing interest.

### Clinical Trial

NOT APPLICABLE

